# Transient peripheral blood transcriptomic response to ketamine treatment in children with ADNP syndrome

**DOI:** 10.1101/2024.01.29.24301949

**Authors:** Ariela S. Buxbaum Grice, Laura Sloofman, Tess Levy, Hannah Walker, Gauri Ganesh, Miguel Rodriguez de los Santos, Pardis Armini, Joseph D. Buxbaum, Alexander Kolevzon, Ana Kostic, Michael S. Breen

## Abstract

Activity-dependent neuroprotective protein (ADNP) syndrome is a rare neurodevelopmental disorder resulting in intellectual disability, developmental delay and autism spectrum disorder (ASD) and is due to mutations in the *ADNP* gene. Ketamine treatment has emerged as a promising therapeutic option for ADNP syndrome, showing safety and apparent behavioral improvements in a first open label study. However, the molecular perturbations induced by ketamine remain poorly understood. Here, we investigated the longitudinal effect of ketamine on the blood transcriptome of 10 individuals with ADNP syndrome. Transcriptomic profiling was performed before and at multiple time points after a single low-dose intravenous ketamine infusion (0.5mg/kg). We show that ketamine triggers immediate and profound gene expression alterations, with specific enrichment of monocyte-related expression patterns. These acute alterations encompass diverse signaling pathways and co-expression networks, implicating up-regulation of immune and inflammatory-related processes and down-regulation of RNA processing mechanisms and metabolism. Notably, these changes exhibit a transient nature, returning to baseline levels 24 hours to 1 week after treatment. These findings enhance our understanding of ketamine’s molecular effects and lay the groundwork for further research elucidating its specific cellular and molecular targets. Moreover, they contribute to the development of therapeutic strategies for ADNP syndrome and potentially, ASD more broadly.

## INTRODUCTION

ADNP syndrome (also known as Helsmoortel-Van Der Aa Syndrome) is a monogenic neurodevelopmental disorder caused by mutations in activity-dependent neuroprotective protein (*ADNP*). ADNP syndrome is clinically characterized by global developmental delay, intellectual disability (ID), and behavioral disorders including autism spectrum disorder (ASD). Although pathogenic mutations in *ADNP* account for ∼0.2% of ASD cases (Gozes, 2020; Siper, et al., 2021; Helsmoortel, et al., 2014), the clinical profile of ADNP syndrome is distinct from individuals with idiopathic ASD (Arnett, et al., 2018); Siper 2021. ADNP syndrome can manifest across an array of physical and psychiatric domains, including motor and language delays, hypotonia, congenital heart disease, sensory reactivity, and additional behavioral manifestations (Helsmoortel, et al., 2014; Siper, et al., 2021). Most infants and children with ADNP syndrome have some form of feeding difficulty, largely caused by dysfunctions in oral motor control (NIH, 2021); apraxia and other motor disorders (Van Dijck, et al., 2019).

ADNP is located on chromosome 20 at q13.13 and encodes for a protein that is crucial for brain function and neurodevelopment (Karmon, et al., 2022; Gozes, 2020). ADNP plays a role in modulating chromatin structure and maintaining gene transcription and neuronal differentiation. *ADNP* interacts directly with two microtubule end-binding proteins (EB1 & EB3) and *ADNP* allelic mutations perturb this interaction (Gozes, et al., 2015; Ivashko-Pachima, et al., 2017). Most pathogenic mutations in *ADNP* involve *de novo* premature termination codons (Chen, et al., 2023) and a loss of function mechanism is therefore assumed. However, it has been shown that mutant transcripts are expressed, consistent with most of the protein being coded from the terminal exon (Gozes, 2020, Breen et al., 2020). The position of the mutation within the ADNP coding sequence results is two very different DNA methylation patterns in blood, which is also not consistent with a simple haploinsufficiency mechanism (Breen et al., 2020).

While there are currently no specific therapeutic strategies for ADNP syndrome, there are studies underway using model systems to assess the efficacy of some targeted therapies. The most extensive research has been on a peptide called NAP, derived from ADNP, where pre-clinical studies in mice suggest a potential for NAP to be protective against damaging mutations in ADNP, but clinical data have not been generated (Gozes, 2020). More recently, ketamine treatment has been proposed as a potential therapeutic for ADNP syndrome. Ketamine is an NMDA receptor agonist with pro-glutamatergic activity, approved for anesthetic purposes and as a therapeutic intervention for treatment-resistant depression (Jia & Hong, 2014; Acevedo-Diaz, et al., 2020). Ketamine’s effects – particularly its adverse events – are generally transient and, while there has been concern about a potential neurotoxic effect of ketamine administered at high doses, (40mg/kg), subclinical and subanesthetic are well tolerated and may, in fact, have neuroprotective effects (Jia & Hong, 2014; Acevedo-Diaz, et al., 2020). To this end, the first open-label safety and preliminary efficacy trial of intravenous ketamine was carried out in ADNP syndrome (Kolevzon, et al., 2022). This study was the first of its class for ADNP syndrome and provides preliminary clinical evidence for improvements in key domains of social communication, attention deficient and hyperactivity, restricted and repetitive behaviors, speech, and activities of daily living.

To build upon these recent efforts, we present complimentary molecular data from individuals with ADNP syndrome who participated in the ketamine trial (Kolevzon, et al., 2022). The primary goal of the current investigation was to examine the transcriptomic response to ketamine treatment in blood and in the immune milieu and to dissect its effects on ADNP-related biology. To do so, we performed a comparative RNA-sequencing analysis of peripheral blood immune cells from 10 individuals with ADNP syndrome before and after a single intravenous infusion of ketamine (0.5mg/kg). A multi-step analytical approach was used that specifically sought to (1) identify individual genes that differ in their response to ketamine, (2) replicate ketamine-induced gene expression profiles in independent transcriptomic studies, and (3) to identify the ketamine-regulated gene networks, thereby providing a functional and mechanistic readout for ketamine response.

## METHODS

### Study design

Peripheral blood biospecimens were collected at the following timepoints: (1) during a screening period within the 4 weeks preceding a baseline visit; (2) immediately following a single low-dose administration of ketamine (0.5mg/kg); and (3) during clinic visits for safety, clinical outcome, and biomarker assessments at day 1, week 1, 2, and 4.

### RNA-sequencing data pre-processing and quality control

Blood was collected in PAXgene tubes and RNA extraction was performed using the PAXgene Blood RNA kit (Qiagen). All samples submitted for sequencing had RNA integrity numbers (RIN) of ≥ 8. RNA-sequencing (RNAseq) was performed by the New York Genome Center in New York, NY, USA. The mRNA TruSeq Stranded kit (Illumina) was used for library preparation and RNA sequencing was run using the NovaSeq system (Illumina) and paired-end chemistry (2x100bp). Notably, PAXgene tubes enable RNA isolation and transcriptome sequencing of peripheral blood leukocytes, which was referred to as the peripheral blood transcriptome in the current study. Sequenced raw RNA reads were aligned to hg38 Ensembl using STAR (Dobin, et al., 2013). RNA quality control was performed using RSeQC (Wang, et al., 2012) and Picard (Broad Institute, 2019) to quantify percent GC, percent duplicates, gene body coverage, and library complexity, as well as to perform unsupervised clustering, and to mark duplicate reads. Next, gene expression was quantified using featureCounts (Liao, et al., 2014). Subsequently, the raw gene expression matrix was filtered to include only genes that were expressed across one-third of the samples in the cohort and normalized using VOOM (Robinson, et al., 2010), resulting in a resulting in a filtered and normalized expression matrix of 17,218 genes.

### Outlier identification and removal

Using the high-quality mapped bam files obtained, we employed the samtools mpileup function to verify the presence of the reported mutation in all individuals (*see details below*). This approach allowed us to confirm the mutation consistently across multiple timepoints for each participant, ensuring reliable validation. We observed two participants (donor IDs: 1668.201, post-infusion timepoint & 1628.201, week 4 timepoint) in whom the expected mutation could not be quantified at one timepoint, differing from the other timepoints. As a result, these two samples were excluded from further analysis since their ADNP mutations could not be accurately quantified and deviated from the remaining repeated measures.

Subsequently, a principal component analysis was conducted on the remaining samples to identify expression outliers that exceeded two standard deviations from the grand mean. Consequently, one sample was considered an outlier and eliminated from the analysis (donor ID: 1329.202, post-infusion timepoint), retaining a total of 51 participants for further analysis.

### Querying ADNP allele-specific expression

The fraction of mutant vs. healthy alleles for the pathogenic ADNP mutation associated with each sample was also quantified, to see whether ADNP allele-specific expression changes corresponded to ketamine treatment. Mapped BAM files from each sample were queried at the location of their associated ADNP mutation site using the mpileup function from Samtools (Danecek, et al., 2021). For samples with a single nucleotide mutation, just the location of the mutated nucleotide was queried; for samples with small deletions or insertions, a region of 10nt, encompassing the affected region, was queried. For mutations affecting two or more nucleotides, the percent reference was calculated as an average of the percent reference alleles across the affected range.

### Cell type deconvolution

CIBERSORTx (https://cibersortx.stanford.edu) was used to perform cell type deconvolution. Cell fractions were imputed using LM22 as the signature matrix (Chen, et al., 2018). Because CIBERSORTx requires gene symbols (as opposed to, for example, EnsIDs), the genes in the unfiltered raw expression matrix were converted to HGNC gene symbols using biomaRt (Durinck, et al., 2005). In instances where two Ensembl IDs corresponded to the same HGNC symbol, the Ensembl ID with highest average expression was retained and the other removed. After conversion to gene symbols, the raw expression matrix contained 40,104 genes, and this matrix was used as the mixture file (without batch correction, and with quantile normalization disabled) for the CIBERSORTx analysis. A total of 15 immune cell types were estimated in this dataset and were subjected to a Dunnett’s test using the DescTools (Signorell & et al, 2022) package to compare baseline cellular proportions (prior to ketamine administration) to subsequent timepoints in the trial.

### Computing gene expression variance explained by technical and biological factors

To determine the best fit model equation for differential gene expression analysis, the variancePartition package (Hoffman & Schadt, 2016) was used. Variance partition analysis employs linear mixed models to quantify the effect of multiple covariates on overall gene expression variability — in this case, subject ID (since each subject was repeated across timepoints), subject sex, subject mutational class (Class I or Class II, see below), subject age, sample RIN, sample timepoint (defined as a factor with six levels: Baseline, Post-infusion, Day 1, Week 1, Week 2, Week 4) were modeled. Here mutational class refer to two classes defined by previously described differences in methylation changes: class I for individuals with mutations located outside a region between nucleotides 2000 and 2340 of the *ADNP* coding sequence and class II for individuals with mutations within this region, including the recurrent mutation p.Tyr719^*^ (Breen et al., 2020; Bend et al., 2019).

### Differential gene expression

Differential gene expression was performed on the VOOM-normalized matrix using the *limma* package (Ritchie, et al., 2015). Differential gene expression covaried for sex as a potential confounding variable as well as and donor as a repeated measure using the duplicateCorrelation() function from limma. A primary linear model was fit to contrast differences during the baseline timepoint versus five post-infusion timepoints (immediately post-infusion, day 1, week 1, week 2, week 4). Differentially expressed genes (DEGs) for each comparison were considered significant if they passed an Benjamini-Hochberg (BH) adjusted p-value threshold of <0.05.

To estimate the influence of varying cell type proportions on differential expression results, estimated cell type proportions, computed via CIBERSORTx, were individually added into each linear model as a covariate. Differential gene expression analysis was performed in the same way and using the same parameters and comparisons as for the original model design. After adjusting for cell type proportion, the number of significant differentially expressed genes from each timepoint relative to baseline (*e*.*g*., post-infusion, day 1, week 1, week 2, week 4) was ascertained. Then, the DEG list from our primary linear model (described above) was intersected with the resulting list of differentially expressed genes with cell type composition as a covariate. The percentage of original DEGs which were no longer significant was computed.

### Querying cell-type specific enrichment of differential expression signatures

A two-step approach was used to dissect the potential impact of cell-type-specific effects on the bulk peripheral blood transcriptome signatures. First, we integrated estimated percent compositions of distinct cellular populations as covariates into the differential expression model equation. This allowed us to compare the DEGs with and without incorporating cell type adjustments, enabling a quantitative assessment of cellular influences on the differential expression comparison. Second, to determine whether the differentially expressed genes or WGCNA modules were associated with any cell type, target gene lists were tested for cell-type specific enrichment, as previously described (Breen et al., 2023). In brief, we leveraged two existing scRNA-seq PBMC datasets, which were generated from two healthy donors. These datasets were processed completely independently — in different laboratories, using different methodologies, and differing in the number of sequenced cells. Both datasets are available as part of 10x Genomic’s publicly accessible data (https://www.10xgenomics.com/resources/datasets). The first dataset, containing 33,227 PBMCs, was processed with Cell Ranger 1.1.0, v.2 Chemistry (10x Genomics, 2016). The second, containing 67,272 PBMCs, was originally published by the Zheng lab (Zheng, et al., 2017) and was processed with Cell Ranger 1.1.0, v.1 Chemistry (10x Genomics, 2016). Each of the two single cell objects had already been filtered, normalized, scaled, and clustered, and the cell type identities assigned.

For each PBMC dataset, cell-type specific enrichment analysis was performed for the following target gene sets: significant DEGs from the Baseline vs. Post-infusion comparison and the Baseline vs. Day 1 comparison. First, the target gene list was intersected with the associated PBMC expression matrix, to generate a PBMC expression matrix of just genes in the target list. The moduleEigengenes function from the WGCNA package was then utilized to summarize each cell’s expression of all the genes in the target list into a single “cell eigenvalue”. These per-cell eigenvalues were then scaled and overlain onto the PBMC UMAP.

### Replication of ketamine-induced transcriptional responses

To replicate the ketamine-induced transcriptomic perturbations, we leveraged two independent transcriptome studies that dissected the impact of ketamine on gene expression profiles (Ho, et al., 2019; Cathomas, et al. 2021).

Ho, et al., 2019 examined the effect of ketamine and its active metabolites on gene expression profiles using the HMC3 human microglial cell line. Here, ketamine, (2*R*,6*R*)-HNK, or (2*S*,6*S*)-HNK (400nM) were administered with or without estradiol (E2; 0.1nM); these concentrations were chosen to emulate observed plasma concentrations seen during ketamine treatment in human subjects (Zarate, et al., 2012; Ho, et al., 2019). To determine the extent of replication, raw FASTQ files for six samples, including two replicates for ketamine and ketamine+E2 treatment, together with two replicates for vehicle alone, were downloaded from GEO (GSE134782).

Cathomas, et al., 2022 explored the effect of intravenous ketamine (0.5mg/kg) on the peripheral blood transcriptome of 26 patients with treatment resistant depression and 21 healthy controls. Notably, blood samples and RNA extraction at baseline and 24 hours post-treatment were ascertained and isolated using similar protocols in the current study. To determine the extent of replication, raw FASTQ files for 42 samples (21 baseline and 21 post-ketamine treatment) were downloaded from GEO (GSE185855).

Data pre-processing of all these two independent transcriptome studies was conducted as described above to ensure accuracy and consistency. Following QC, RNA-sequencing mapping and counting, raw expression matrices were filtered and normalized, and samples were clustered using PCA and MDs plots. To generate ketamine-induced transcriptional responses from each of these studies, we performed the following analyses.

Due to sample size limitations for Ho et al., 2019, we computed log_2_ fold-changes for each condition (ketamine and ketamine + E2) relative to vehicle. Thus, a log_2_ fold-change was generated for each gene specific to each treatment condition. For Cathomas, et al., 2022, we used the following model in the limma R package : (∼0 + *Time* + *Group* + *Age*), in which Time was a factor reflecting baseline or post-ketamine treatment, Age was the patient age, and Group was a factor representing responder or non-responder status (responders were originally defined by a ≥50% reduction of baseline MADRS score). Donor as a repeated measure was also accounted for using *duplicateCorrelation() from the limma R package*. Subsequently, log_2_ fold-changes were leveraged for comparative purposes.

Next, to determine transcriptome-wide concordance of ketamine-induced gene expression responses across all comparisons, we evaluated the correlation coefficients between log_2_ fold-change values across the entire transcriptome as a measure of replication. For comparative purposes, we used only genes that were in common with pass filter genes in the current study (12040 genes in common for Ho, et al., and 13532 genes in common for Cathomas, et al.)

### Weighted gene co-expression network analysis

Weighted gene co-expression network analysis (WGCNA) (Langfelder & Horvath, 2008) was performed on the VOOM normalized expression matrix of 17,218 genes across all timepoints jointly. The co-expression similarity matrix was generated; a soft thresholding power of 11 was selected to be applied to the co-expression similarity matrix to calculate adjacencies. The signed (values ranging from -1 to 1) adjacency matrix was then used to determine similarity — or overlap — by generating the Topological Overlap Matrix (TOM). The gene dendrogram was generated by performing average linkage hierarchical clustering on the gene expression dissimilarities (1 - *TOMsimilarity*), so that the distance between any two groups was defined as the average of all pair-wise distances of the genes within those two groups. Both hybrid and dynamic cuts were plotted to determine the best method for identifying gene clusters. Ultimately, the gene dendrogram was generated using a cut height of 0.99999999999, and modules were identified using the dynamic cut method with a deep split value of 1, with a minimum cluster size (number of genes in the module) of 50. A resulting 34 gene modules were identified for downstream analysis.

Each module’s first principal component was used to calculate the module eigengene — a single value summarizing a modules’ gene expression. The Pearson’s correlations between module eigengenes (MEs) and several subject and sample traits were calculated, including: subject sex, subject mutational class (Class I or Class II), sample timepoint (defined as a factor with levels: Baseline, Post-infusion, Day 1, Week 1, Week 2, Week 4), and sample cell type composition (defined as a continuous variable for the percent composition of each of the identified cell types). In addition to using embedded WGCNA tools to determine whether modules were significantly associated with time, a Dunnett’s test was also run on the module MEs to see whether any modules were significantly different between timepoint comparisons.

### Functional annotation of differentially expressed genes and co-expression modules

Functional annotation of identified DEGs and co-expression modules was run using the online portal offered by ToppGene (https://toppgene.cchmc.org). For each of the five timepoint comparisons, two DEG lists were generated (one of significant upregulated genes and one of significant downregulated genes, as described in *2*.*3*). These lists were then submitted as the Training set for ToppGene: Candidate gene prioritization. Training parameters were set for: 1) FDR correction, 2) a p-value (determined by the probability density function) cutoff of 0.05, 3) gene limits 1 ≤ n ≤ 2000. Test parameters were set for a random sampling size of 5000, and a minimum feature count of 2. ToppGene IDs from the following selected features: GO: Molecular Function, GO: Biological Process, GO: Cellular Component, Human Phenotype, Pathway, Gene Family, Drug, and Disease were used downstream for REVIGO analysis (http://revigo.irb.hr).

Protein–protein interactions were obtained from the STRING database (Szklarczyk et al., 2019) with a signature query of gene co-expression modules identified from our analysis. STRING implements a scoring scheme to report the confidence level for each direct protein–protein interaction (low confidence: <0.4; medium: 0.4–0.7; high: >0.7). We used a combined STRING score of >0.4. Hub genes within the protein–protein interaction network is defined as genes with the highest degree of network connections.

### Enrichment analysis of targeted gene sets within co-expression modules

All co-expression modules identified by WGCNA analysis were tested for targeted gene set enrichment using GeneOverlap (Shen, 2022). This function uses a Fisher’s exact test and an estimated odds-ratio for all pair-wise tests based on a background set of genes detected in the current study. Enrichment was queried for the following targeted gene sets: 1) significant DEGs observed in the current study; 2) curated lists of ASD risk genes taken from two published reports (Satterstrom et al., 2020 & Fu et al., 2022); and 3) curated list of ADNP-interacting or -associated genes based on multiple lines of independent evidence (BioGRID (https://www.pathwaycommons.org), SFARI (https://gene.sfari.org), and INNATEDB (Lynn, et al., 2008; Breuer, et al., 2013; Lynn, et al., 2010)).

## RESULTS

### ADNP allele-specific expression patterns

The present cohort is comprised of 10 children with ADNP syndrome (7 male, 3 female) (**Supplemental Table 1**). The peripheral blood transcriptome was systematically profiled at six distinct timepoints for each participant (**Figure 1A**). Participants harbored nine unique pathogenic variants within *ADNP*, thereby capturing the genetic heterogeneity associated with this syndrome (**Figure 1B**). Among these variants, six were classified as frameshift mutations, and three were identified as nonsense mutations, with two participants exhibiting the recurrent p.Tyr719* variant.

**Figure 1.**
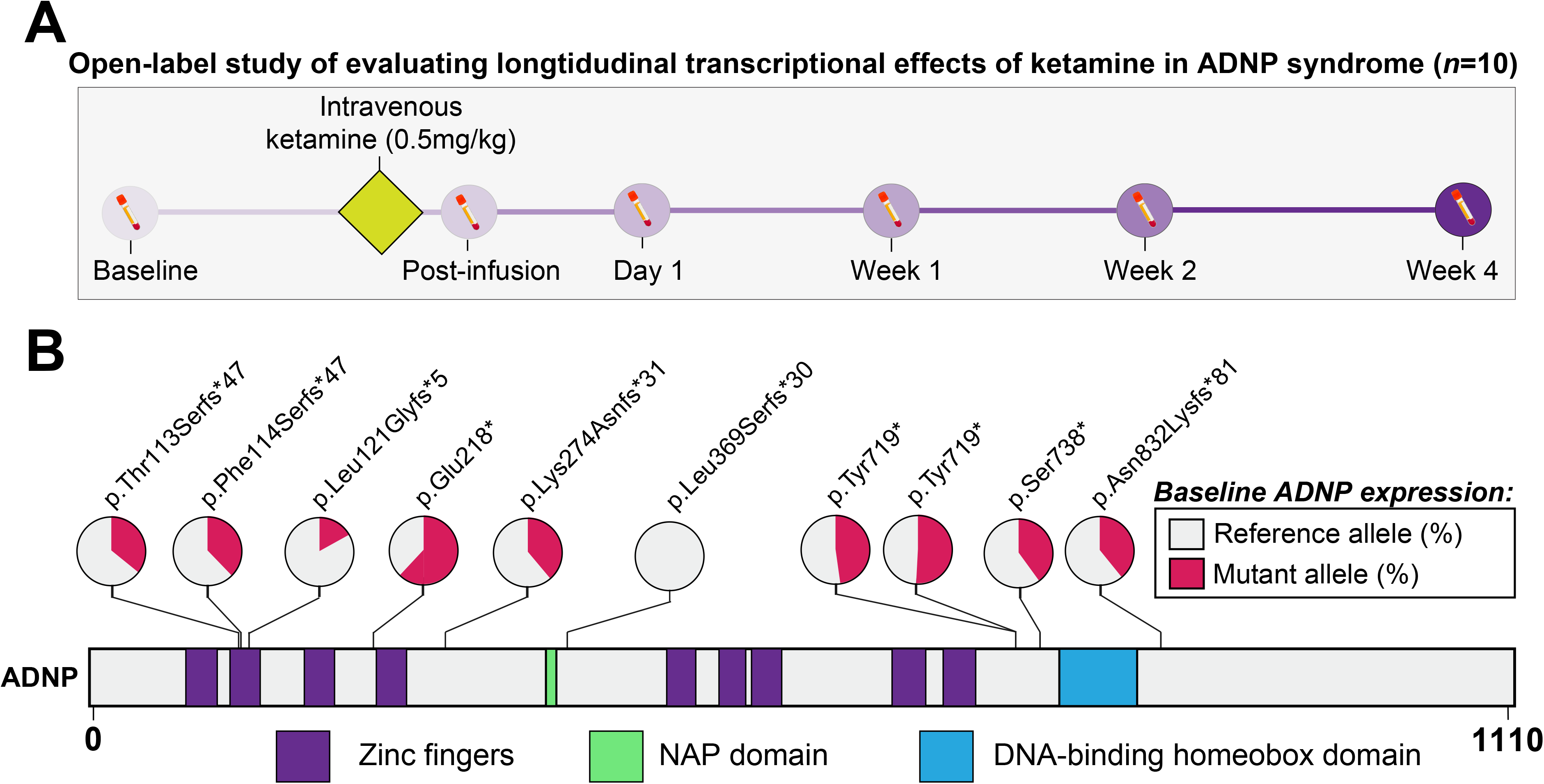
Trial design and *ADNP* allele-specific expression. (**A**) Clinical trial workflow and data collected across the time course. A total of 10 pediatric participants were enrolled in the trial and were screened within 28 days of the trial onset. Post-infusion represents the same day as intravenous ketamine treatment (0.5 mg/kg, over 40 minutes), while Day 1 is the following day. (**B**) Pathogenic variants represented by the study cohort and their locations along the ADNP gene locus. Two patients had recurrent p.Tyr719* mutations. *ADNP* RNA-seq read pileups were queried for each mutation (57.66 ± 22.35 read coverage). The percent of *ADNP* reference (healthy) allele at baseline (or at 4 weeks, if baseline measurements were not available) are in grey, while the percent of *ADNP* mutant allele is in red.

While *ADNP* mutations have been thought to result in haploinsufficiency, recent investigations have revealed that mutant *ADNP* alleles are not subject to nonsense-mediated decay and are associated with distinct methylation changes in blood. Thus, for each participant in this cohort, we performed a targeted analysis and queried the expression of ANDP mutant and reference (healthy) alleles (*see Materials and Methods*). Presence of *ADNP* mutant allele was confirmed in 9 out of 10 participants, whereby reference alleles ranged from 49%-83% of total ADNP expression (mean 61%; **Figure 1B**). Notably, the ratio of reference over mutant allele expression for ADNP remained consistent throughout the entire trial across all participants (**Supplemental Figure 1A, Supplemental Table 1**). Likewise, the gene expression profile for *ADNP* mRNA did not exhibit statistically significant changes in response to ketamine treatment, nor did the expression of other known ASD-related genes (**Supplemental Figure 1B & 1C**).

### Early immediate transcriptomic response to ketamine treatment

To comprehensively assess the transcriptome-wide effects of ketamine treatment, we first evaluated the impact of various factors on gene expression profiles. For each gene, a linear mixed model was employed to determine the proportion of gene expression variation attributable to different biological and technical covariates (**Supplemental Figure 2**). Collectively, these covariates accounted for approximately 48% of the mean transcriptome variation. Among them, donor treated as a repeated measure exerted the most substantial genome-wide effect, explaining a median of 28% of the observed variation. Additionally, timepoint and RNA integrity numbers (RIN) contributed to transcriptome variability, explaining approximately 3.7% and 2.7%, respectively. The remaining factors accounted for less than 1% of the overall transcriptome variability. Subsequently, to determine differential gene expression, we conducted an analysis comparing gene expression levels at each timepoint to the baseline, while accounting for potential confounding effects of donor as a repeated measure, RIN, and biological sex. This analysis identified ketamine-induced differentially expressed genes (DEGs; FDR < 5%): 1325 DEGs at post-infusion (689 upregulated, 636 downregulated); 776 DEGs at day 1 (227 upregulated, 549 downregulated); 122 DEGs at week 2 (18 upregulated, 104 downregulated); 80 DEGs at week 4 (10 upregulated, 70 downregulated) (**Figure 2A, Supplemental Table 2**). No significant DEGs were identified at week 1. Overall, the strongest transcriptional effect was observed immediately after ketamine treatment at the post-infusion timepoint (**Figure 2B**), and most of these genes were not dysregulated at later timepoints (**Supplemental Figure 3)**, indicating that their expression patterns resumed baseline expression levels one day later.

**Figure 2.**
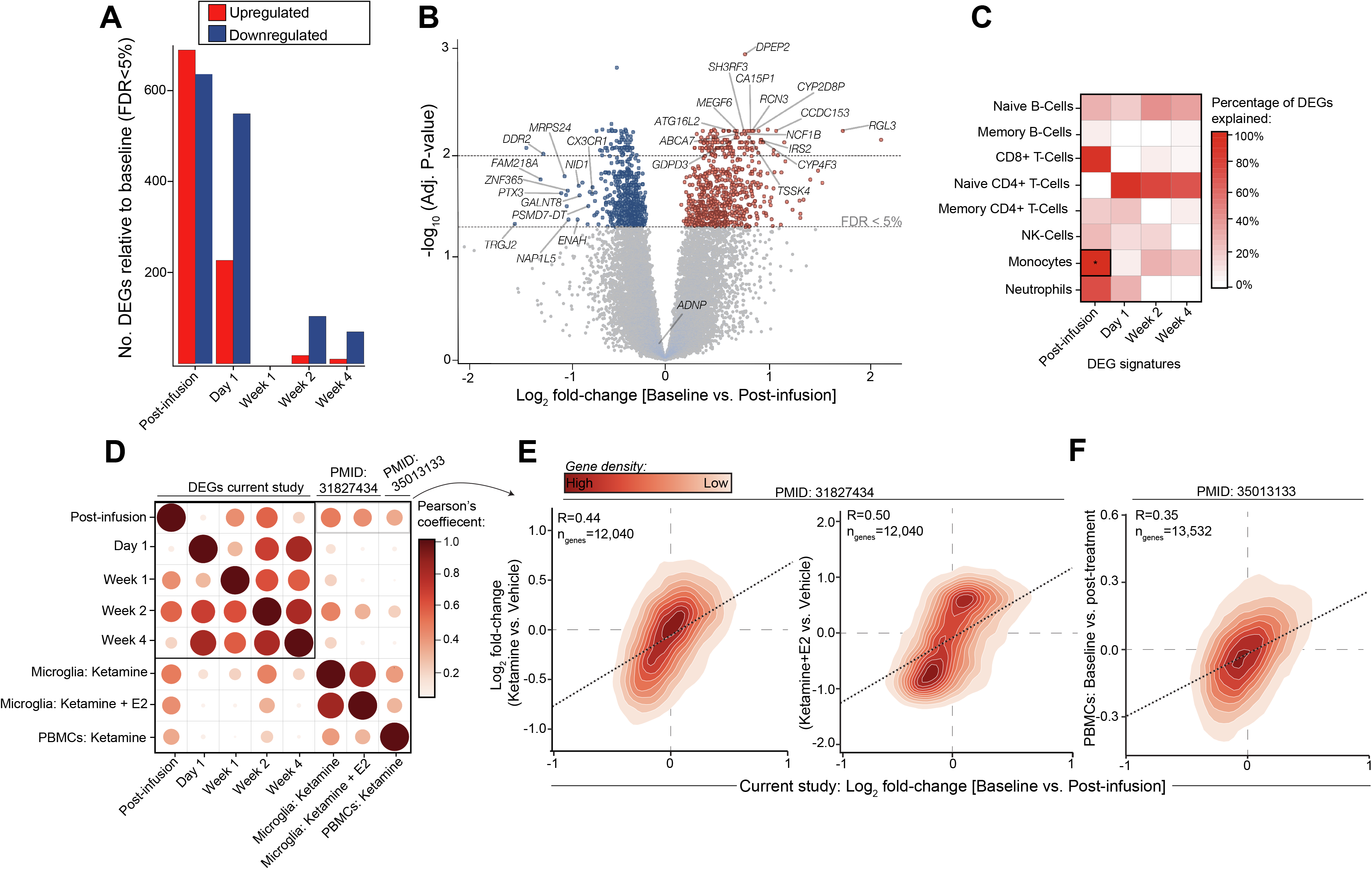
Ketamine-induced gene expression perturbations in peripheral blood cells. (**A**) The number of differentially expressed genes (DEGs) passing FDR < 5% (x-axis) at each timepoint compared to baseline (y-axis). Upregulated genes are shown in red and downregulated genes are shown in blue. (**B**) Volcano plot (x-axis: logFC, y-axis: -log10(p.adj)) illustrating DEGs identified in the baseline vs. post-infusion comparison, with colored points representing genes passing FDR < 5%. (**C**) Heatmap of DEGs shared before and after covarying for cell type (y-axis) at each timepoint comparison (x-axis). Covarying for monocytes showed the strongest effect when comparing baseline and post-infusion, accounting for more than 98% of the observed DEG signal. (**D**) Correlation plot of transcriptome-wide log2 fold-changes of differentially expressed genes identified at each timepoint, compared to results from a study in immortalized microglia (Ho et al.) (*Microglia: Ketamine*, ketamine vs. vehicle; *Microglia: Ketamine + E2*, ketamine and estradiol vs. vehicle) and a study of baseline vs. post-treatment expression in a cohort of individuals with treatment-resistant depression (Cathomas et al.) (*PBMCs: Ketamine)*. Circle size and color (scaled from 0-1) represent correlation strength. Density plots comparing baseline vs. post-infusion from the current study (x-axis) relative to (**E**) findings from microglia treated with ketamine (left) and ketamine + estradiol [E2] (right) (y-axis) and relative to (**F**) PBMCs from individuals treated with a single-low dose ketamine infusion (y-axis). R values of the lm fit values are indicated above the regression lines. PMIDs are provided for each independent study, respectively.

### Post-infusion differential expression is explained by monocyte-specific gene signatures

Cell type proportions were included as covariates into our linear models to gauge their influence on differential gene expression signatures. We observed pronounced differences on the post-infusion DEG signature when accounting for monocytes (**Figure 2C**), which contributed to more than 98% of the observed DEG profile. Notably, the estimated proportion of circulating monocytes significantly decreased post-infusion (Dunnett’s test, *p*-value= 0.047, ∼4.5% change), whereas the estimated frequencies for all other cell types were stable and did not change over the time course (**Supplemental Figure 4, Supplemental Table 2**). Next, to support this observation, we queried the expression of the post-infusion DEGs within thousands of single peripheral blood mononuclear cells across two independent scRNA-seq experiments (*see Methods*). These analyses confirmed that post-infusion DEGs are preferentially and highly expressed in CD14+ monocytes (**Supplemental Figure 5**). Importantly, such enrichment was not observed in other cell type populations, nor for any other DEG signature from other timepoints, suggesting that acute effects of ketamine may perturb monocyte-specific gene expression.

### Replicating ketamine-induced gene expression profiles in independent transcriptomic studies

To validate the observed ketamine-induced gene expression signatures, we turned to two independent transcriptomic studies that investigate the impact of ketamine. In the first study, the HMC3 human microglial cell line was used to examine the effect of ketamine or ketamine with estradiol (E2) relative to vehicle conditions. The second study explored the impact of intravenous ketamine (0.5mg/kg) on the peripheral blood transcriptome of individuals with treatment resistant depression, specifically at 24 hours post-treatment, relative to an untreated baseline timepoint. To ensure accuracy and comparability, all data from these studies were downloaded, re-processed and analyzed using the same methodology (*see Methods*). To determine replication of ketamine-induced transcriptomic signatures, we evaluated the transcriptome-wide concordance of ketamine-induced log2 fold-changes across all comparisons in the current study alongside the findings from the two independent reports (**Figure 2D**). Encouragingly, a high level of concordance was observed across the different time points in the current study. Notably, the post-infusion timepoint exhibited substantial transcriptome-wide concordance not only with ketamine-induced effects in microglia (ketamine, *R*=0.48; ketamine+E2 *R*=0.54) (**Figure 2E**), but also with ketamine-induced effects in peripheral blood (*R*=0.35) (**Figure 2F**). These results indicate that the early immediate transcriptomic response to ketamine that we observed in ADNP syndrome replicate across different studies and models.

### Co-expression network analysis informs ketamine-induced gene networks

To gain further biological insights to the ketamine-induced gene expression patterns, we employed WGCNA. This approach aggregates gene expression variation into discrete co-expression modules across. Through this analysis, we identified 34 co-expression modules (**Figure 3A; Supplemental Table 3**) varying in size from 51 to 1356 genes. Of these modules, six exhibited significant changes in expression levels relative to baseline at one or more subsequent time points (Dunnett’s test, *p < 0*.*05*) (**Figure 3B**). For instance, modules M1 (232 genes) and M2 (735 genes) displayed significant down-regulation post-infusion compared to baseline (*p*=1.93x10^−8^, *p*=5.20x10^−70^, respectively) and these modules were strongly enriched for functions related to mRNA processing and RNA metabolism (**Figure 3C**). Conversely, modules M3 (96 genes) and M4 (1040 genes) were significantly up-regulated at post-infusion relative to baseline (*p*=5.45x10^−24^, *p*=3.01x10^−93^, respectively). Module M4 was significantly enriched for immune and inflammatory responses and MAPK cascades (**Figure 3C, Supplemental Figure 6A)**. In addition, down-regulation of module M5 (557 genes) at day 1 (*p*=7.15x10^−95^) was observed and enriched for processes in the blood and cellular components such as platelets and plasma, as well as processes involving myeloid cells (**Supplemental Figure 6B**). Finally, up-regulation of module M6 (55 genes) was observed post-infusion and day 1 (*p*=7.82x10^−11^, *p*=3.73x10^−2^, respectively), however did not show significant functional annotation.

**Figure 3.**
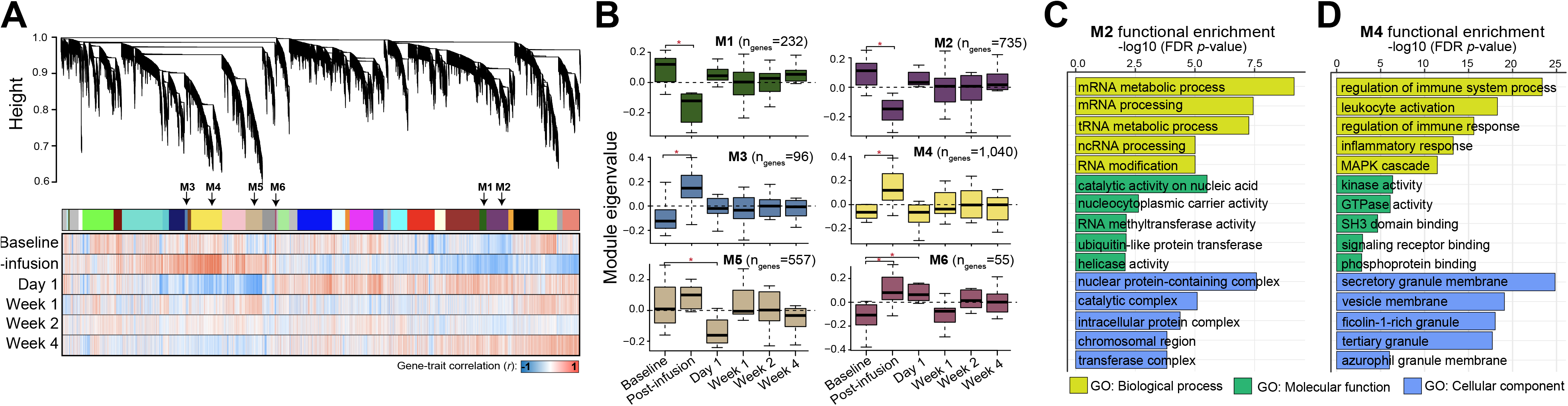
Ketamine-induced changes in gene co-expression patterns. (**A**) Gene dendrogram (top) of WCGNA analysis applied to the VOOM normalized expression matrix of 17,218 genes across all timepoints jointly. A total of 34 co-expression modules were identified using the dynamic split method colored (upper bar). Each line on the dendrogram represents a single gene. Color bars indicate the association of each gene with the respective timepoint (negative correlations in blue, positive correlations in red). (**B**) Boxplots of the ME values (y-axis) of the six modules showing significant association (Dunnett’s test, *p* < 0.05) with time at one or more timepoints (x-axis). (**C & D**) GO Functional enrichment plots of the purple (M2; left) and yellow (M4; right) modules, which reflect the two modules with the strongest signal for functional enrichment. Bar color indicates GO functional group, and each bar is labelled with the associated enrichment term. Top significant terms are shown for each functional group, with the -log_10_ of the multiple test corrected p-values (x-axis) shown for each enrichment term (y-axis).

To further explore the potential involvement of ASD risk genes, we curated well-known lists of ASD-associated genes and examined their co-expression and dynamic ketamine-induced transcriptional responses in peripheral blood. However, no significant enrichment was observed, aligning with our previous analysis (**Supplemental Figure 7**). Notably, we also did not observe enrichment for curated lists of ADNP interacting genes within any co-expression modules in the current study (**Supplemental Figure 7**). Overall, these findings indicate that ketamine treatment exerts a transient effect characterized by the up-regulation of inflammatory responses and the down-regulation of RNA processes, which quickly return to baseline levels within one day. These responses do not include ADNP nor a significant proportion of other ASD risk genes.

## DISCUSSION

This study investigated the transcriptomic response to ketamine treatment in children with ADNP syndrome, a monogenic neurodevelopmental disorder associated with ASD. Our findings contribute to the understanding of ketamine’s effects on gene expression and provide insights into potential therapeutic mechanisms for ketamine on behavioral health and on ADNP syndrome. We explore three therapeutic mechanisms for ketamine including one hypothesis-driven and two unbiased approaches: (1) effects of ketamine on the expression of ADNP and other ASD-associated genes; (2) acute effects of ketamine on transcriptome-wide and gene module expression; and (3) persistent effects of ketamine on transcriptome-wide and gene module expression.

One mechanism we exclude is a direct effect of ketamine treatment on the expression levels of *ADNP* or other known ASD-related genes. This observation aligns with independent investigations that also show limited alterations in ASD-related gene expression following ketamine administration (Ho, et al., 2019; Cathomas, et al. 2021). As such, clinical benefits observed with ketamine are likely mediated by broader downstream effects of ketamine, such as modulation of synaptic plasticity, glutamate signaling, or neuroinflammatory responses (Nugent, et al., 2019; Kopelman, et al., 2023; Abdallah, et al., 2018; Li, et al., 2020; Chen, et al., 2021; Grieco, et al., 2021; Chen, et al., 2018).

The transcriptome-wide response to ketamine treatment exhibited dynamic patterns across different timepoints, providing valuable insights into the temporal dynamics of ketamine-induced gene expression changes. The most pronounced transcriptional effect was observed immediately following ketamine infusion, characterized by a substantial number of DEGs. However, most of these genes returned to baseline expression levels within one day, providing the opportunity to examine acute and persistent effects of ketamine on gene expression. The acute findings are in line with previous research highlighting the acute and short-lasting effects of ketamine on neuronal activity and synaptic plasticity (Kim, et al., 2022; Chen, et al., 2021; Li, et al., 2010; Kopelman, et al., 2023; Grieco, et al., 2021). The rapid normalization of gene expression suggests that ketamine-induced alterations in transcription are part of a dynamic and regulated process, potentially involving the transient modulation of signaling pathways or feedback mechanisms. By integrating estimated cell-type compositions, our analysis revealed that the post-infusion DEG signature was primarily driven by changes in monocyte-specific gene expression. Monocytes, as immune cells involved in inflammatory responses, play crucial roles in shaping the immune milieu within the central nervous system (Peng, et al., 2022; Arteaga-Henríquez, et al., 2022; Xie, et al., 2017). The upregulation of genes associated with immune and inflammatory processes observed in our study aligns with previous evidence highlighting the involvement of the immune system in the pathophysiology of neurodevelopmental disorders, including ASD (Arteaga-Henríquez, et al., 2022l; Molloy, et al., 2005; Gupta, et al., 1998; Dantzer, et al., 2008). Moreover, the enrichment of post-infusion DEGs in monocytes, as confirmed by single-cell RNA sequencing experiments, provides further support for the pivotal role of monocyte-driven transcriptomic alterations in response to ketamine treatment. CD14+ monocytes, known as a subset of monocytes with distinct functional properties, have been implicated in neuroinflammation and have been shown to contribute to the pathogenesis of neurodegenerative disorders and psychiatric conditions (Padmos, et al., 2008; Nowak, et al., 2019; Rodríguez, et al., 2017). The preferential expression of ketamine-induced DEGs within CD14+ monocytes suggests their active participation in mediating the transcriptomic response to ketamine administration and points towards the potential involvement of neuroinflammatory processes in the therapeutic effects of ketamine.

To validate our findings and assess the generalizability of ketamine-induced gene expression signatures, we compared our acute results with two independent transcriptomic studies investigating the effects of ketamine. Encouragingly, we observed a high level of concordance between the transcriptome-wide effects of ketamine in ADNP syndrome and the effects observed in microglia and peripheral blood of individuals with treatment-resistant depression. This replication across different experimental and clinical models supports the notion that the molecular pathways and mechanisms underlying the therapeutic effects of ketamine exhibit a degree of consistency and universality across various neurological and psychiatric conditions. Microglia, the resident immune cells of the central nervous system, and monocytes, a subset of immune cells found in peripheral blood, share common developmental origins and functional properties. Both cell types play crucial roles in immune responses and inflammation within the brain (Andoh, 2021; Ritzel, et al., 2015). The fact that ketamine-induced gene expression changes in ADNP syndrome show concordance with those observed in microglia and peripheral blood further supports the involvement of immune-related processes in the therapeutic effects of ketamine.

We performed weighted gene co-expression network analysis to further explore the biological processes associated with ketamine treatment in ADNP syndrome. Notably, modules associated with mRNA processing and RNA metabolism exhibited down-regulation, suggesting a potential dampening effect of ketamine on these processes. This observation aligns with previous studies reporting the acute effects of ketamine on neuronal activity and synaptic plasticity, which involve the regulation of RNA processes (Ho, et al., 2019; Kim, et al., 2022; Cathomas, et al., 2022). In contrast, modules related to immune and inflammatory responses displayed significant up-regulation following ketamine treatment. These findings support the notion that ketamine exerts immunomodulatory effects, consistent with emerging evidence indicating the involvement of the immune system in the pathophysiology of neurodevelopmental disorders (Arteaga-Henríquez, et al., 2022; Molloy, et al., 2005; Tanabe, et al, 2018). The up regulation of genes associated with immune and inflammatory processes highlights the potential role of ketamine in modulating inflammatory pathways in ADNP syndrome. Interestingly, the observed changes in gene expression were transient, with most genes returning to baseline expression levels within one day post-infusion. This suggests that ketamine elicits a transient perturbation of molecular pathways, which subsequently return to their basal state. This dynamic response may reflect the rapid adaptation of cellular processes following ketamine administration, potentially involving feedback mechanisms and homeostatic regulation.

Beyond the acute effects, unraveling ketamine’s sustained effects on gene expression is crucial for gaining insights into its prolonged therapeutic potential and potential adverse effects, particularly in the context of treating ADNP syndrome, diverse neuropsychiatric disorders, and chronic pain. Towards this end, we noted that ketamine exerted lasting effects on a select set of genes, particularly those encompassed in co-expression module M6 (**Figure 3**). Although the functional annotation of these genes still remains elusive, a number of individual genes with sustained effects were notable. Of special interest are several metabolism-related genes that showed upregulation in response to ketamine. These include genes involved in cellular metabolism (e.g. *NADSYN1*), cholesterol metabolism (e.g. *LCAT* and *SOAT1*), lipid metabolism (e.g. *ACOT8*), and fatty acid metabolism (e.g. *CPT1A*) (Natesan & Kim, 2021). While there isn’t a direct established connection with ketamine, the role of cholesterol metabolism and lipid signaling in neurological disorders has been previously indicated, thus making it an intriguing domain for mental health research. Another gene of interest is *SCN9A*, which encodes for a voltage-gated sodium channel involved in pain perception (Cox et al., 2006). Although ketamine is acknowledged for its pain-modulating properties, its association with *SCN9A* remains largely unexplored. Overall, it is vital to continue expanding our knowledge of ketamine’s long-term effects on gene expression, a research area that holds promise in improving our understanding of ketamine’s complex mechanism of action. This, in turn, will aid in the design and development of newer, more effective, and safer medications derived from or inspired by ketamine.

It is also important to acknowledge several limitations that should be considered when interpreting the results of this study. Firstly, the sample size of the cohort with ADNP syndrome included in this study was relatively small, which may limit the generalizability of the findings to the broader ADNP syndrome population. Although we were able to demonstrate the generalizability of ketamine-induced transcriptomic signatures across independent studies, future investigations with larger cohorts are needed to validate and expand upon these results. Secondly, it is important to recognize that the effects of ketamine treatment can vary depending on various factors, including individual patient characteristics. Although these factors were held constant in our study, they may influence the observed gene expression changes and should be considered when interpreting the results. Thirdly, while we employed a two-step analytical approach and utilized orthogonal datasets to assess the cellular specificity of the post-infusion DEG signal, further studies focusing on high-dimensional immune profiling or fluorescence-activated cell sorting could provide a more precise measurement of cell type fluctuations following treatment. Lastly, although we employed a rigorous methodology to evaluate the peripheral blood transcriptome, it is essential to acknowledge that blood-based gene expression profiles may not fully capture the complexity of gene expression patterns in specific brain regions or cell types that may be more directly relevant to the pathophysiology of ADNP syndrome. Therefore, caution should be exercised when extrapolating these findings to brain-specific mechanisms.

This study provides valuable insights into the transcriptomic response to ketamine treatment in individuals with ADNP syndrome. The dynamic and time-dependent nature of the response highlights the complex molecular mechanisms underlying ketamine’s therapeutic actions. The prominent role of monocyte-specific gene expression alterations suggests a potential link between immune and inflammatory processes and the therapeutic effects of ketamine in neurodevelopmental disorders. The down-regulation of mRNA processing and RNA metabolism modules, coupled with the up-regulation of immune and inflammatory response modules, underscores the multifaceted nature of ketamine’s molecular effects. These findings deepen our understanding of the intricate pathways affected by ketamine and provide a foundation for future research aimed at unraveling the precise cellular and molecular targets of ketamine treatment. Moving forward, it is essential to conduct further investigations to elucidate the specific molecular pathways and downstream effects of ketamine treatment in ADNP syndrome. Additionally, future research endeavors should encompass larger cohorts, repeated dosing, experimentally tractable neuronal model systems, longer follow-up periods, and diverse populations to validate the generalizability and clinical implications of our findings.

## Data Availability

RNA-sequencing fastq files have been deposited in the Gene Expression Omnibus under accession number GSE235921.

## Acknowledgements

We are extremely grateful to the families who participated and to the ADNP Kids Research Foundation and the Foundation for Mood Disorders for their support. Support for mediKanren was provided by the National Center for Advancing Translational Sciences, National Institutes of Health, through the Biomedical Data Translator Program, awards OT2TR003435 and OT2TR002517. Any opinions expressed in this document are those of the Translator community at large and do not necessarily reflect the views of NCATS, individual Translator team members, or affiliated organizations and institutions. ClinicalTrials.gov Identifier: NCT04388774, Low-Dose Ketamine in Children with ADNP Syndrome, https://clinicaltrials.gov/ct2/show/NCT04388774.

## Data and code availability

RNA-sequencing fastq files have been deposited in the Gene Expression Omnibus under accession number GSEXXXXX (data will be uploaded prior to date of publication). Computational code and intermediate data files required for reproducibility of these results is available on GitHub (https://github.com/buxgrice/adnpket).

## Ethics and approval to participate

All study participants have given written informed consent, and the genetic study has been approved by the Icahn School of Medicine at Mount Sinai Institutional Review Board.

## Competing interests

Alexander Kolevzon is on the scientific advisory boards of Ovid Therapeutics, Ritrova Therapeutics, and Jaguar Therapeutics and consults to Acadia, Alkermes, GW Pharmaceuticals, Neuren Pharmaceuticals, Clinilabs Drug Development Corporation, and Scioto Biosciences. Joseph D. Buxbaum consults for BridgeBio Pharma. The remainder of the authors declare no competing interests.

## Notes

### Clinical Trial

NCT04388774

