## Supplemental figures for "Transient peripheral blood transcriptomic response to ketamine treatment in children with ADNP syndrome"

This file contains seven supplemental figures:

**Supplemental Figure 1. Gene expression of ADNP and other autism risk genes.**
**Supplemental Figure 2. Variance partition showing the variance explained by covariates.**
**Supplemental Figure 3. Differentially expressed genes between other trial timepoints and baseline.**
**Supplemental Figure 4. Estimated cell-type composition across trial timepoints.**
**Supplemental Figure 5. Enrichment of differentially expressed genes in independent scRNA-seq data.**
**Supplemental Figure 6. Protein-interaction networks of candidate modules.**
**Supplemental Figure 7. Enrichment of candidate gene sets in co-expression modules.**

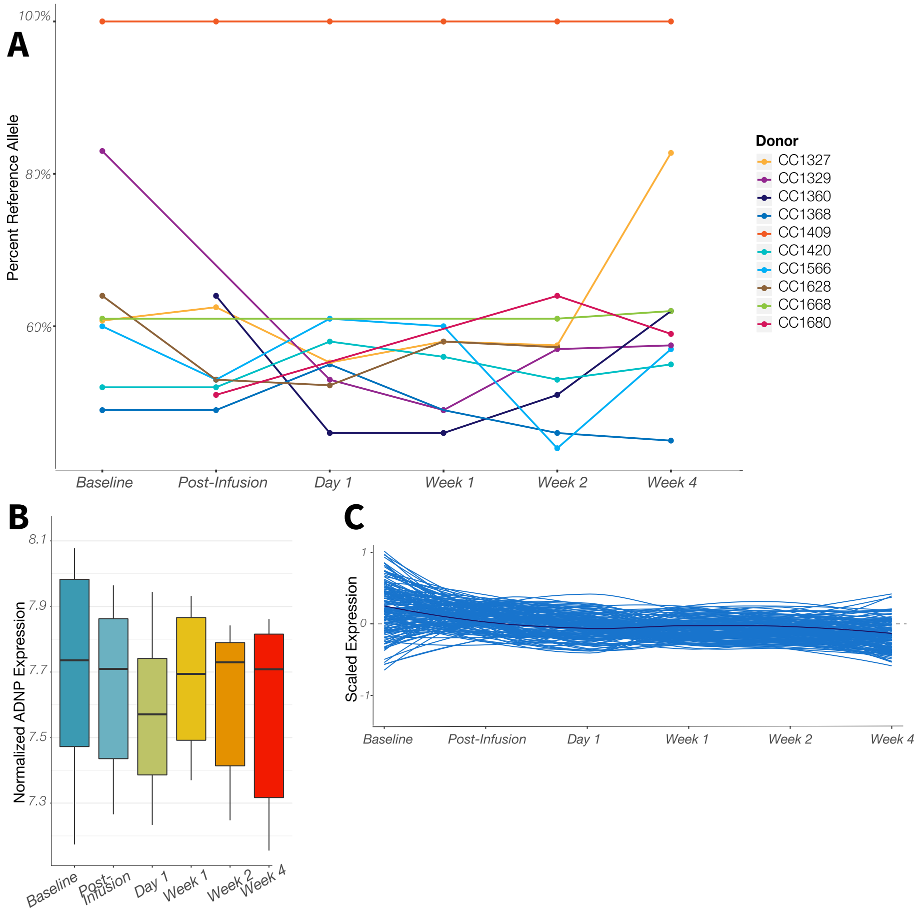


**Supplemental Figure 1. Gene expression of ADNP and other autism risk genes.** (**A**) Percent of reference (healthy) allele (y-axis) across trial timepoints (x-axis) for 10 pediatric cases of ADNP Syndrome included in this study. Samples that were removed in QC steps are not included. Subject CC1409 did not express the mutant allele in blood data (p.Leu369Serfs*30). (**B**) Normalized expression of *ADNP* (log2 counts per million) (y-axis) from participants across trial timepoints (x-axis). (**C**) Z-scaled expression of 151 of the 183 ASD risk genes from Fu, et al., (2021) expressed in the normalized gene expression matrix. *ADNP* is colored in dark blue.

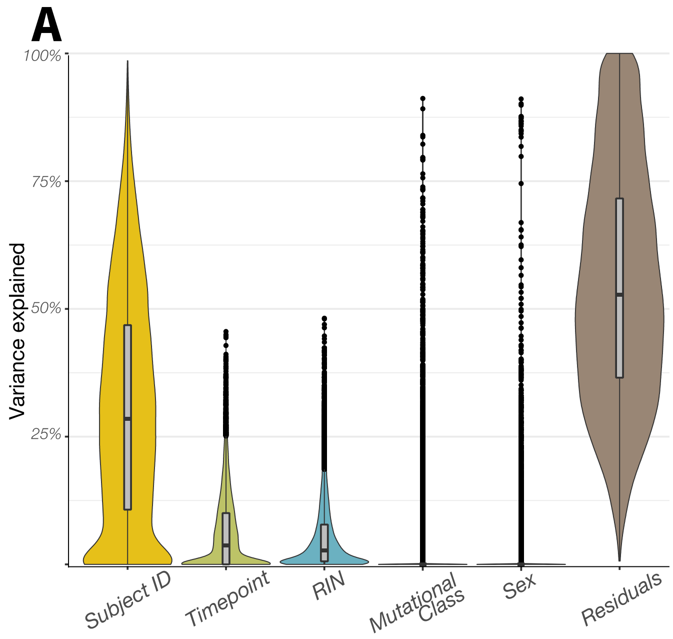


**Supplemental Figure 2. Variance partition showing the variance explained by covariates.** (**A**) Variance partition plot showing the percentage of transcriptomic variance (y-axis) explained by covariates in the current study (x-axis), including donor as a repeated measure, timepoint, RNA integrity numbers (RIN), mutational class, sex and residual variation.

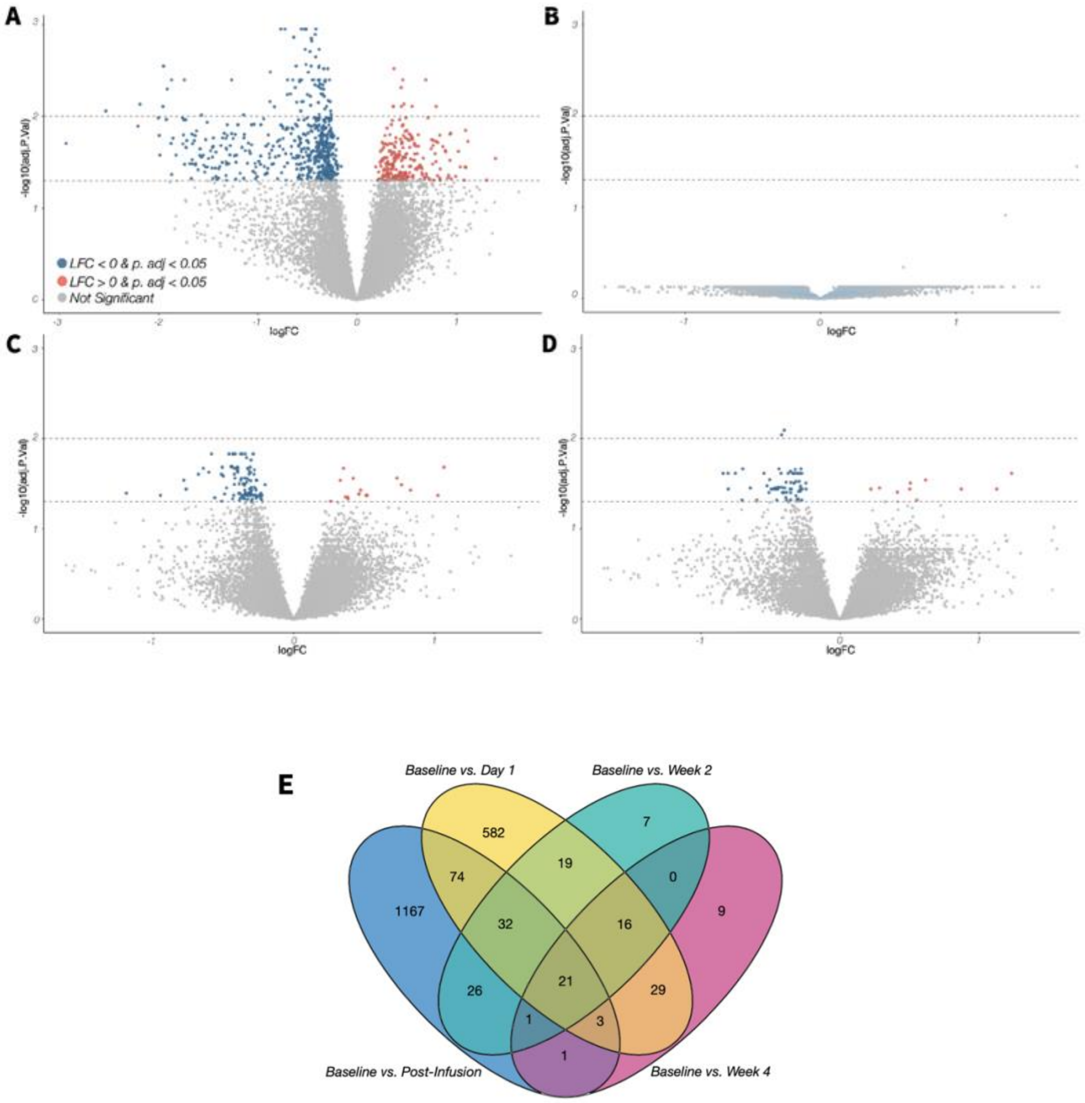


**Supplemental Figure 3. Differentially expressed genes between other trial timepoints and baseline.** Volcano plots of differentially expressed genes (DEGs) at post-treatment timepoints: (**A**) Day 1 vs. Baseline; (**B**) Week 1 vs. Baseline; (**C**) Week 2 vs. Baseline; (**D**) Week 4 vs. Baseline. Log2 fold-change (logFC, x-axis) relative to -$\log_{10}$ adjusted *p*-value (y-axis). Genes passing a p.adj. < 0.05 are in blue if downregulated and red if upregulated. (**E**) Venn diagram representing the number of significantly DEGs (p.adj < 0.05) at each comparison (Baseline vs. Week 1 had no significant DEGs) and the number of intersecting genes between pairwise and multi-comparison lists. In blue is Baseline vs. Post-Infusion; in yellow is Baseline vs. Day 1; in teal is Baseline vs. Week 2; in pink is Baseline vs. Week 4.

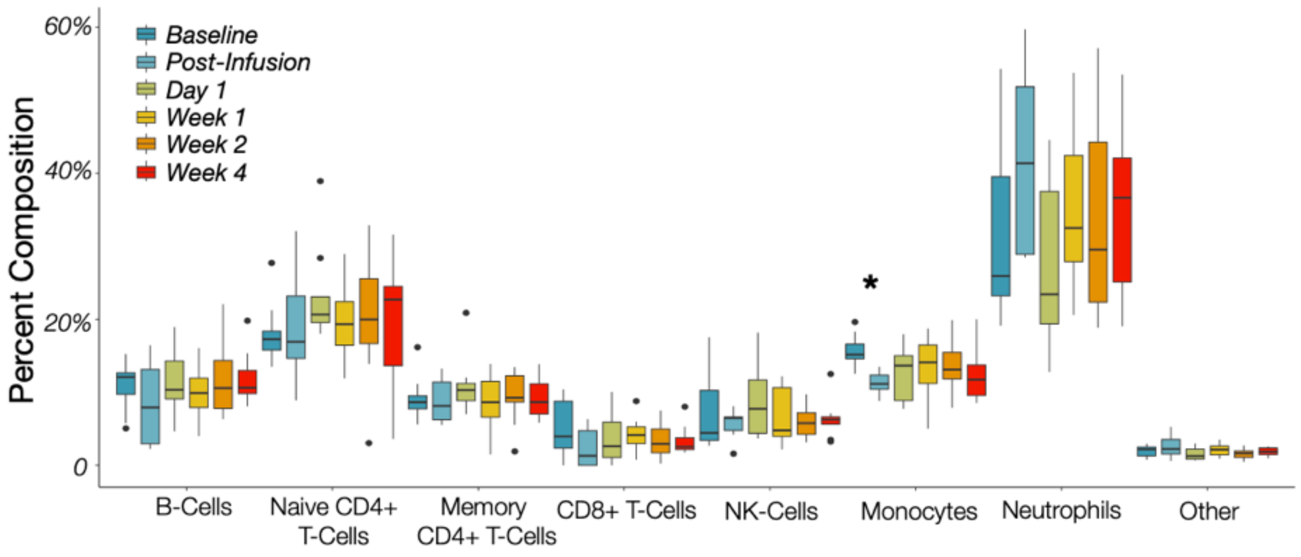


**Supplemental Figure 4. Estimated cell-type composition across trial timepoints.** The estimated percentage of CIBERSORTx cellular composition (y-axis) for each major cell population (x-axis) by trial timepoint. Box color corresponds to trial timepoint. Some cell types are summarized into one larger group: "*B-Cells*" is the summed percent of naïve and memory B-Cells; "*Memory CD4+ T-Cells*" is the summed percent of resting and activated memory CD4+ T-Cells; "*NK-Cells*" is the summed percent of resting and activated NK-Cells; "*Other*" is the summed percent of low frequency M0 Macrophages, activated dendritic cells, and resting mast cells. A Dunnett's test was used to test for significant ($p < 0.05$) differences between baseline composition and post-treatment timepoints; the only comparison reaching significance (starred) is the estimated percent composition of monocytes at the post-infusion timepoint compared to baseline.

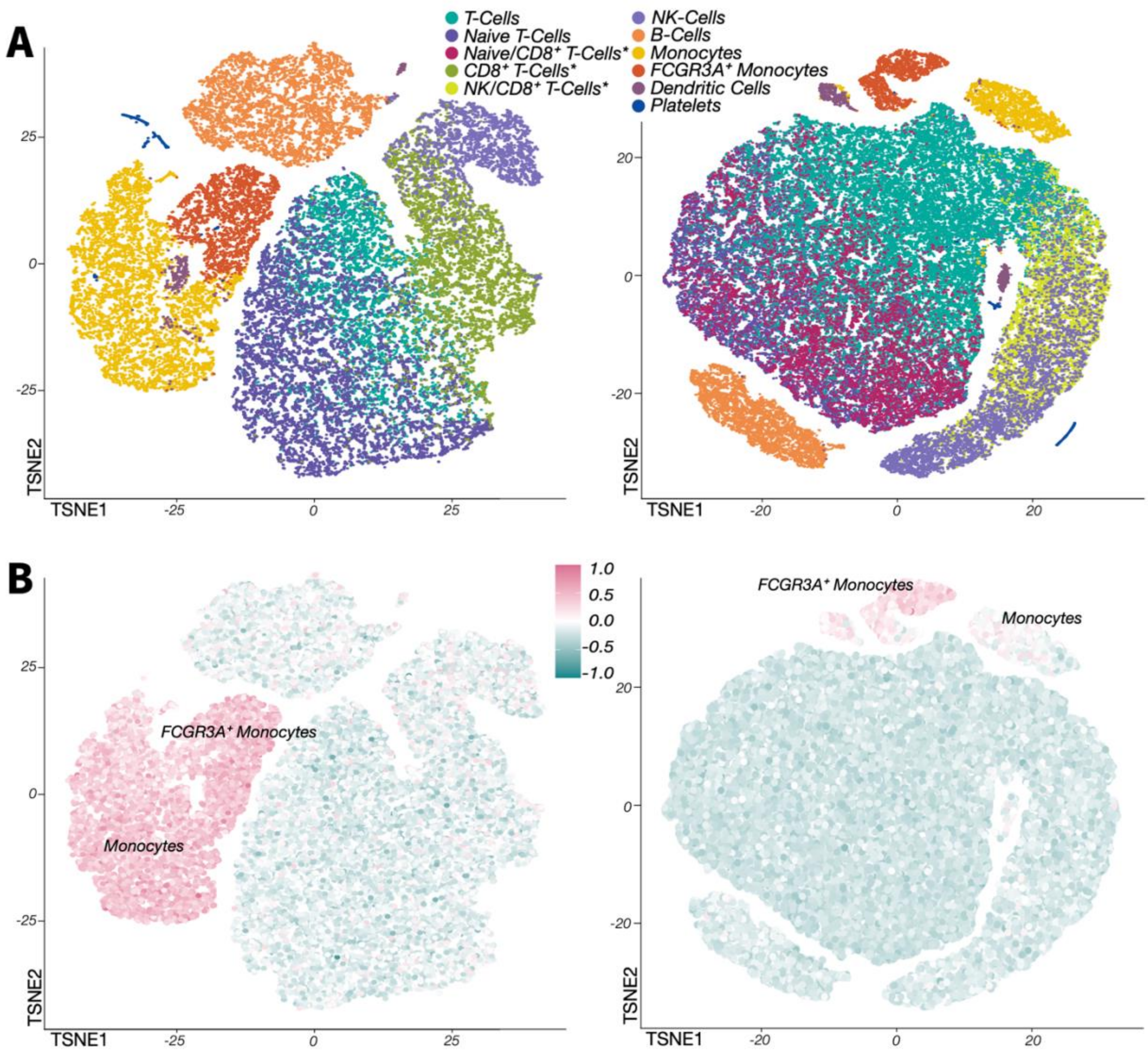


**Supplemental Figure 5. Enrichment of differentially expressed genes in independent scRNA-seq data.** (**A**) Dimensional reduction of two sets of PBMCs from unaffiliated healthy donors, comprised of 33k (left) or 68k (right) cells. Cell type annotations were made using canonical marker genes and are colored as shown. Cell types with a * next to them are groups only identified in one of the two PBMC sets. (**B**) Enrichment of significant ($p < 0.05$) DEGs from the *Baseline vs. Post-Infusion* comparison in the PBMC sets. The color of each cell represents a single value (eigenvalue) of the relative enrichment of the 1325 genes in the DEG list in that cell's gene expression profile. Eigenvalue ranges for both PBMC sets are rescaled to range from 1 to -1, with 1 (pink) representing stronger enrichment and -1 (turquoise) representing weaker enrichment. Monocytes and FCGR3A+ Monocytes are labelled.

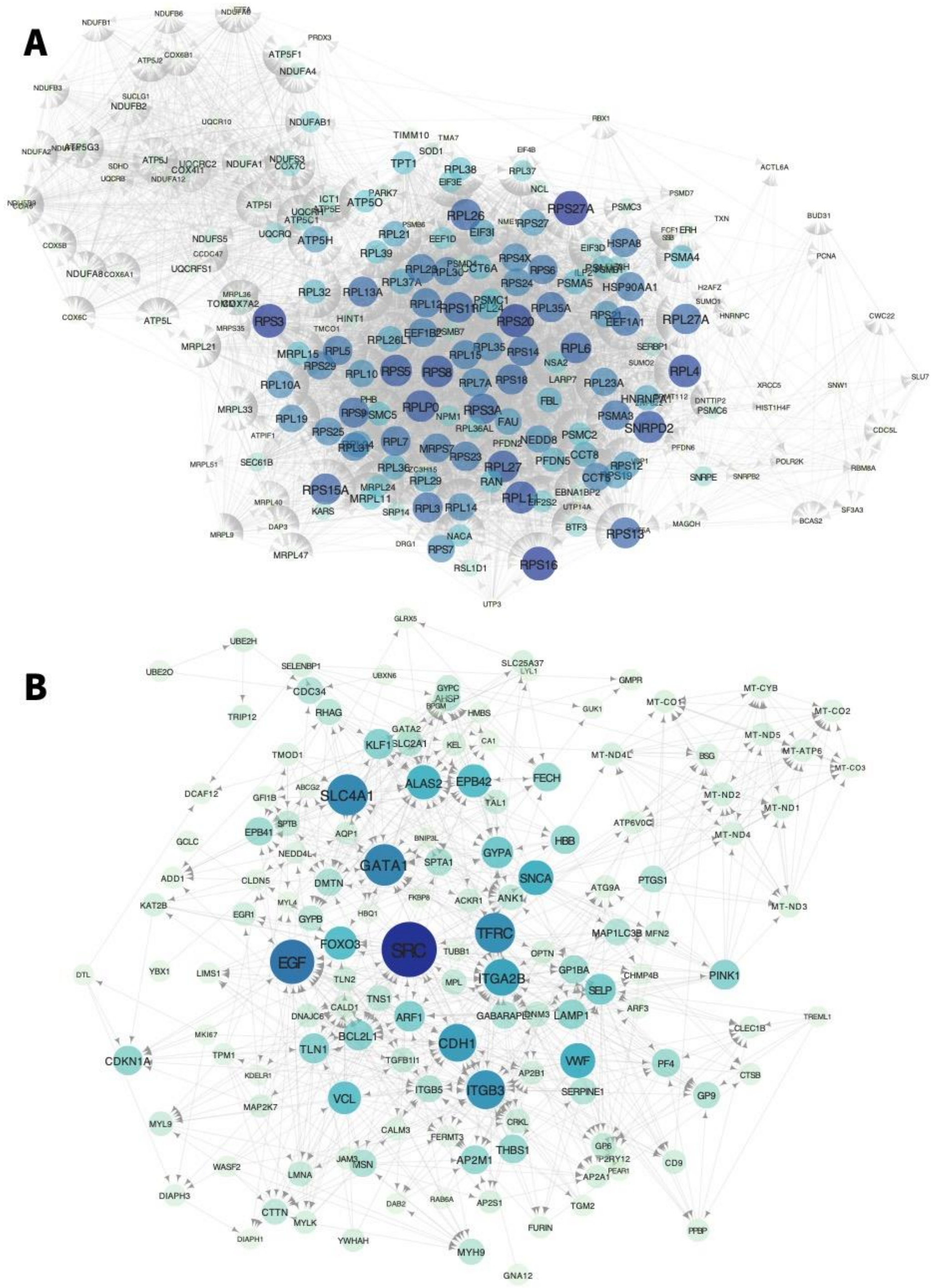


**Supplemental Figure 6. Protein-interaction networks of candidate modules** Genes in the yellow module (M4) and the tan module (M6) were analyzed in STRING to generate a protein-protein interaction network, which was then graphed in Cytoscape. Node color (yellow to blue) and label size are scaled with the number of edges for each node, such that nodes with more edges are larger and colored blue, and nodes with few edges are small and yellow. (**A**) Gene interaction network for the yellow module. Nodes are additionally filtered to include only those with an edge count of between 31 and 143 (the maximum), inclusive OR an edge combined score of between 0.62 and 0.999, inclusive. (**B**) Gene interaction network for the tan module. Nodes are additionally filtered to include only those with an edge count of between 10 and 72 (the maximum), inclusive OR an edge combined score of between 0.475 and 0.999, inclusive.

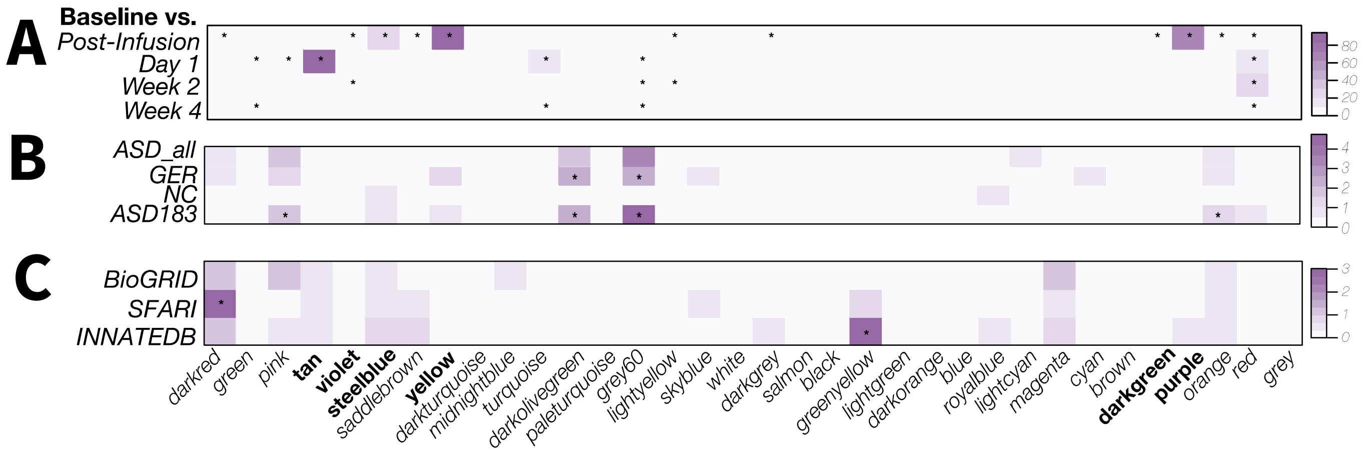


**Supplemental Figure 7. Enrichment of candidate gene sets in co-expression modules.** Targeted enrichment tests of the 34 modules identified by WGCNA analysis, with module colors on the x-axis (bottom) and the modules significantly associated with time in bold. Color intensity scales with strength of enrichment between genes in each module and the targeted gene set (y-axis) and significant ($p < 0.05$) enrichment is annotated with a star. (**A**) Enrichment of lists of significant DEGs (passing *p.adj* threshold of 0.05) from each post-treatment timepoint versus baseline in WGCNA modules. (**B**) Enrichment of three curated lists of autism risk genes (bars 2-4) and a merged "master list" (bar 1) in WGCNA modules. (**C**) Enrichment of generated lists of ADNP-interacting genes from 3 sources (BioGRID, SFARI, and INNATEDB) in WGCNA modules.
